# Performance comparison of ten state-of-the-art machine learning algorithms for outcome prediction modeling of radiation-induced toxicity

**DOI:** 10.1101/2024.05.24.24307747

**Authors:** Ramon M. Salazar, Saurabh S. Nair, Alexandra O. Leone, Ting Xu, Raymond P. Mumme, Jack D. Duryea, Brian De, Kelsey L. Corrigan, Michael K. Rooney, Matthew S. Ning, Prajnan Das, Emma B. Holliday, Zhongxing Liao, Laurence E. Court, Joshua S. Niedzielski

## Abstract

**Purpose:** To evaluate the efficacy of prominent machine learning algorithms in predicting normal tissue complication probability utilizing clinical data obtained from two distinct disease sites, and to create a software tool that facilitates the automatic determination of the optimal algorithm to model any given labeled dataset.

**Methods and Materials:** We obtained 3 sets of radiation toxicity data (478 patients) from our clinic, gastrointestinal toxicity (GIT), radiation pneumonitis (RP), and radiation esophagitis (RE). These data comprised clinicopathological and dosimetric information for patients diagnosed with non-small cell lung cancer and anal squamous cell carcinoma. Each dataset was modeled using ten commonly employed machine learning algorithms (elastic net, LASSO, random forest, regression forest, support vector machine, XGBoost, k-nearest-neighbors, neural network, Bayesian-LASSO, and Bayesian neural network) by randomly dividing the dataset into a training and test set. The training set was used to create and tune the model, and the test set served to assess it by calculating performance metrics. This process was repeated 100 times by each algorithm for each dataset. Figures were generated to visually compare the performance of the algorithms. A graphical user interface was developed to automate this whole process.

**Results:** LASSO achieved the highest area under the precision-recall curve (AUPRC) (0.807±0.067) for RE, random forest for GIT (0.726±0.096), and the neural network for RP (0.878±0.060). Area-under-the-curve was 0.754±0.069, 0.889±0.043, and 0.905±0.045, respectively. The graphical user interface was used to compare all algorithms for each dataset automatically. When averaging AUPRC across all toxicities, Bayesian-LASSO was the best model.

**Conclusion:** Our results show that there is no best algorithm for all datasets. Therefore, it is important to compare multiple algorithms when training an outcome prediction model on a new dataset. The graphical user interface created for this study automatically compares the performance of these ten algorithms for any dataset.

## INTRODUCTION

In radiation oncology, the development of outcome prediction models offers an objective approach to personalized cancer treatment [1]. These models, based on machine learning (ML), provide a data-driven understanding of patient-specific responses to radiation therapy, thereby enabling clinicians to anticipate and mitigate potential toxicities more accurately [2–4]. This enhances the precision of therapeutic interventions and may significantly improve patient outcomes by preemptively addressing the likelihood of specific toxicities.

Outcome prediction models (OPMs) of radiation pneumonitis (RP) from radiation therapy of non-small cell lung cancer (NSCLC) can identify patients with a high risk of developing this toxicity based on their treatment plan, which provides information on the likelihood of lung toxicity when adjusting radiation dose [5]. Similar models may provide predictions for the likelihood of acute radiation esophagitis (RE), a dose-limiting toxicity that can greatly reduce patient quality of life [6]. This may inform potential early plan adaptation strategies to minimize the probability of RE [7]. For pelvic cancers, OPMs are valuable tools in assessing the risk of gastrointestinal toxicities (GIT), which may help guide the choice of radiation techniques to minimize damage to the gastrointestinal tract [8]. These are some examples of how OPMs can form part of a clinical decision support system, enhancing patient care by creating opportunities to reduce the severity of toxicities.

The advancement of OPMs, however, faces some challenges. One of them is the lack of comparative analyses of different ML algorithms tailored for radiation-induced toxicity prediction. The landscape of predictive models is greatly varied, with new types of algorithms being constantly introduced, and there exists no universally acknowledged best algorithm which suits all datasets [9]. Given this diversity, researchers must rely on their personal experience, ease of implementation, or prevalence in the literature to select an algorithm, yet most existing studies focus on a single algorithm for a specific disease site, limiting our understanding of how different models perform across various types of toxicities.

Another critical issue is the interpretability of OPMs. Many advanced algorithms lack transparency in their decision-making processes. In a field where clinical decision-making is highly nuanced and patient-specific, the inability to interpret and understand the rationale behind model-based predictions is a significant drawback. This lack of transparency can impede the trust and adoption of OPMs by radiation oncology professionals [10].

Previous work by Deist et al. has shown that there is no algorithm type which is superior in performance across all datasets [11]. This highlights the need for a comparative multi-algorithm approach to outcome prediction. A free, open-source package which automatically conducts these analyses can address this need, while also enabling a wider range of healthcare institutions to benefit from cutting-edge ML technologies. Such a package could foster collaborative improvement and validation of the algorithms, as the global medical physics community could contribute to its refinement and customization [12].

While some free, open-source packages offering automatic multi-algorithm comparisons do exist, they are designed as general-purpose tools for virtually any ML task [13]. However, it has been shown that, while these tools may show superior performance in some domains, they may underperform in others [14]. Hence, the development of a specialized package for multi-algorithm analysis tailored for radiation therapy is essential.

In our study, we develop a graphical user interface (GUI) using the open-source R package caret (version 6.0.90) to facilitate and automate the comparison of multiple models for any radiation therapy dataset [15,16]. We use this GUI to compare the efficacy of different OPMs in predicting radiotherapy outcomes. We seek to evaluate a variety of state-of-the-art algorithms across three radiation-induced toxicities to investigate if their performance is dataset dependent. The GUI provides mathematically robust interpretations for each model and a systematic way to select the best model for each application. This analysis is essential for researchers seeking a rigorous method of model selection. The code for this GUI will be made available upon publication of this study.

## MATERIAL AND METHODS

### Patient Characteristics

We obtained a waiver of consent and approval from the institutional review board for this retrospective study. Three toxicity datasets (478 patients in total) were collected from our institution. One dataset involved 246 SCCA (squamous cell carcinoma of the anus) patients with different grades of acute GIT (Suppl. Table S1) [17]. The second dataset consisted of 232 NSCLC (non-small cell lung cancer) patients, where the outcome of interest was RP (Suppl. Table S2). The third dataset contained the same 232 NSCLC patients, but the outcome of interest was RE (Suppl. Table S3).

The 246 patients with SCCA were treated from 2003 to 2019 with definitive intensity-modulated radiotherapy (IMRT) or volumetric modulated arc therapy (VMAT) based chemoradiotherapy (CRT) at our institution. All of them were treated in the head-first to gantry, supine position, and had at least 3 months of clinical follow up. They all received conventionally fractionated radiation using a simultaneous integrated boost technique; most patients received 2 Gy per fraction to the primary tumor to a total dose of 50-58 Gy and 1.6-1.7 Gy per fraction to the elective nodal volume to a total dose of 43-47 Gy. Additionally, 236 patients underwent concurrent chemotherapy, 6 received sequential chemotherapy, and 4 received no chemotherapy.

The 232 patients with NSCLC were treated between 2006 and 2019 at our institution with either intensity modulated radiation therapy (IMRT/VMAT, n = 116), passive-scatter proton therapy (PSPT, n = 71), or intensity-modulated proton therapy (IMPT, n =45). The IMRT/VMAT treatment plans were designed using Pinnacle (Philips healthcare; Cambridge, MA) and proton treatments were planned using Eclipse (Varian Medical Systems; Palo Alto, CA). All patients received fractionated radiation consisting of doses between 1.8–3.0 Gy per fraction.

Total dose ranged from 60–74 Gy and patients were treated 5 times per week. All patients were treated with either induction, concurrent, or adjuvant chemotherapy in addition to RT.

### Toxicity Evaluation

GITs were graded by the treating physician during weekly see and follow up visits according to the National Cancer Institute’s Common Terminology Criteria for Adverse Events (CTCAE) version 5.0 and include: abdominal pain, colitis, colonic disorders, constipation, diarrhea, enterocolitis, fecal incontinence, lower GI hemorrhage, nausea, malabsorption, small intestine disorders, and vomiting [18]. All these toxicities were reported to be grade 0 for all patients at the start of treatment. The highest grade on any of the listed disorders within 3 to 6 months of treatment was recorded as the toxicity outcome for a patient. The outcomes were then divided into two classes, with one class being a toxicity less than grade 3, and the other being a toxicity greater than or equal to grade 3. The percentage of patients who developed a grade 3 or higher acute GIT was 9.5% (N=28).

Radiation pneumonitis was graded from 0 to 5 according to the CTCAE v5.0 [18]. The approximate time for RP to develop in a patient after radiation therapy is between 1 to 6 months [19]. The highest grade within two years of treatment was recorded as the toxicity outcome for that patient for this study. A patient with an RP grade ≥2 was considered symptomatic for RP and hence the outcomes were separated into two classes, with one being RP grade ≥2 and the other being a RP grade<2. The percentage of patients who developed a grade 2 or higher for RP was 29.3% (N=68).

Similarly, RE was also graded from 0 to 5 according to CTCAE v5.0 [18]. The highest grade within two years of treatment was recorded as the toxicity outcome for that patient for this study. However, RE peaked during treatment. A patient with an RE grade ≥2 was considered symptomatic for RE, and hence patients with an RE grade ≥2 were classed separate from those with an RE grade<2. The percentage of patients who developed a grade 2 or higher for RE was 60% (N=139).

### Dose-Volume Histogram Metrics

Dose-volume histogram (DVH) metrics were extracted from the Raystation treatment planning system. The metrics extracted were the mean, maximum, and minimum dose, the relative volume receiving at least a given dose (i.e. the value obtained for V30Gy[%] refers to the percent volume of a structure receiving at least 30 Gy) from 5–50 Gy in 5 Gy increments, and the total volume of the region of interest (the full bowel bag for GIT, the lung volume minus the tumor volume for RP, and the esophagus for RE).

### Algorithms

Ten different algorithms were chosen, based on their frequent usage in medical data analysis, to model the three toxicities. They are briefly described below, but more information can be found about them in the literature [20]:

### Model Construction

The schematic of model construction is illustrated in Fig. 1. Each dataset was partitioned into a test set and a training set. Training/test set splits of 0.5, 0.6, 0.7, 0.8, and 0.9 were analyzed. An 8-times repeated 2-fold cross-validation was used on the SCCA training dataset for tuning the hyperparameters of each individual model type and each dataset split. For the NSCLC datasets, a 6-times repeated 3-fold cross-validation was implemented instead. All model types were run in regression mode except for random forest, which was run in classification mode. Hyperparameter tuning was achieved by minimizing the root mean square error for the regression models, and by maximizing kappa for the classifier. Models were trained on the training set and assessed on the test set to compute their performance metrics. The dataset split with the best final performance based on the area under the precision-recall curve was selected as the final model for each algorithm type.

**Figure 1.**
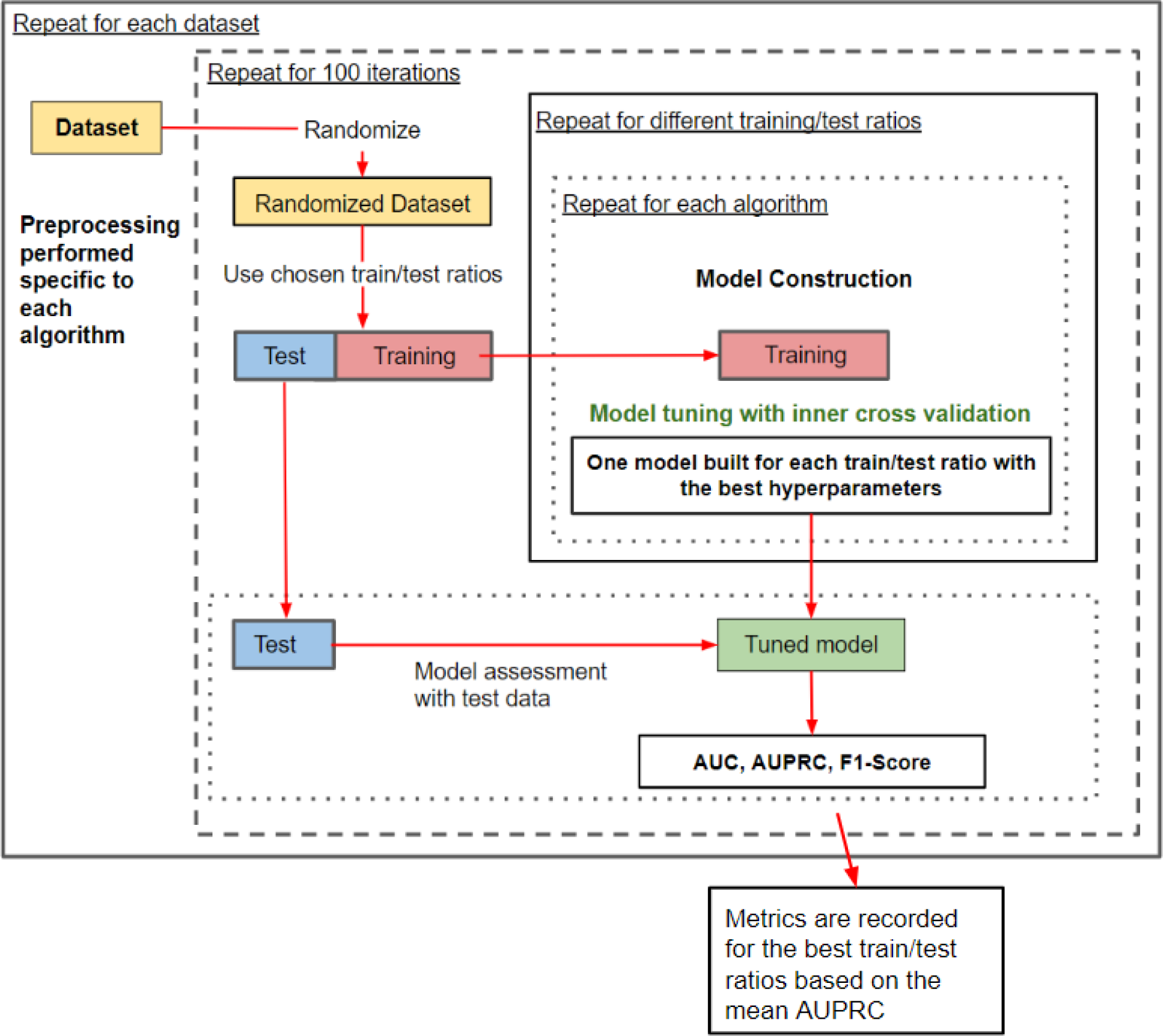
Diagram of the model building and assessment process. Preprocessing, depending on the model being trained, may involve dummy coding, deleting zero variance features, or rescaling. The randomization and splitting of the initial dataset correspond to a Monte Carlo cross-validation approach for the outer loop. The inner loop undergoes repeated k-fold cross-validation or out-of-bag error minimization for model construction.

This whole procedure was repeated 100 times with different randomization seeds.

However, it is important to note that the same set of 100 randomization seeds was used for the 100 iterations of each algorithm type and dataset split such that, for the same iteration number, the models were trained and tested on identical training and test sets for a specific dataset. This allows for the performance metrics of any two algorithms to be analyzed through a pairwise comparison. The evaluation metrics calculated for each algorithm are accuracy, area under the receiver operating characteristic curve (AUC), area under the precision-recall curve (AUPRC), and the F1-score.

Note that, for the algorithms trained in regression mode, the output they provide for each patient is a numerical score ranging from 0 to 1. This score is proportional to the probability of developing the toxicity in question. Accuracy is calculated by setting a threshold of 0.5, where patients with a score above the threshold are considered to be at risk of developing the toxicity. AUC and AUPRC are evaluated by varying the risk threshold from 0 to 1, plotting its effect on sensitivity/specificity or precision/recall, respectively, and then finding the area under the curve. The F1-score is measured at an optimal threshold that will maximize precision and recall when assigning equal importance to both metrics.

Algorithm rankings were computed for each dataset and iteration by arranging the AUPRC or AUC for each model type in descending order. For simplicity, we limited the comparative analysis to AUC and AUPRC results.

We compared the effectiveness of choosing an algorithm based on its performance during the training cross-validation phase (dataset specific selection) against random model selection (which mimics a choice without prior modeling knowledge or dataset consideration). Changes in predictive performance between those selection criteria were measured.

### Graphical User Interface

To facilitate the training and comparison of these algorithms, we created a graphical user interface (GUI) using the shiny library in R. This tool automates the process of model training, cross-validation, and performance analysis for a given dataset. It eliminates the need to develop expertise in the ten algorithms discussed here before being able to select one adequately.

The GUI allows the user to input any data as a csv file. Once loaded, the user will see a list of all the features in the data file. Checkboxes will be available to select only the desired features. A drop-down menu is also available for designating the outcome variable. A report for exploratory data analysis may also be automatically generated (Suppl. Fig. S1).

Next, one can select which algorithms will be trained, which training/test split ratios will be analyzed, how many iterations should be performed, what kind of cross-validation procedure should be used (Monte Carlo, k-fold, repeated k-fold, or leave-one-out), and whether an automatic feature selection (based on permutation importance) should be conducted. Recommended default values are provided for convenience (Suppl. Fig. S2).

Any missing values in both training and test sets are imputed using medians for continuous attributes and modes for categorical attributes from the training set. Categorical attributes are subjected to dummy coding when required, which expresses categorical features through a set of binary attributes. Covariates with zero variance or near-zero variance are eliminated when necessary. Training and test set features are re-scaled and normalized.

Hyperparameter tuning is also performed automatically. A random search is initially performed across the hyperparameter phase space. The optimal result of that search is used as a central point for a grid search around that neighborhood. After this grid search, another random search is performed. If the results of this random search are better than those of the grid search, the process is repeated. Otherwise, the process ends, and the hyperparameters found using the grid search are kept.

After training the models, their individual accuracy, AUC, AUPRC, and F1-score are calculated. Plots and tables are generated to help visualize the difference between the performances of these algorithms (Figs. 2-6). The user should then be able to select the best algorithm for their application.

**Figure 2.**
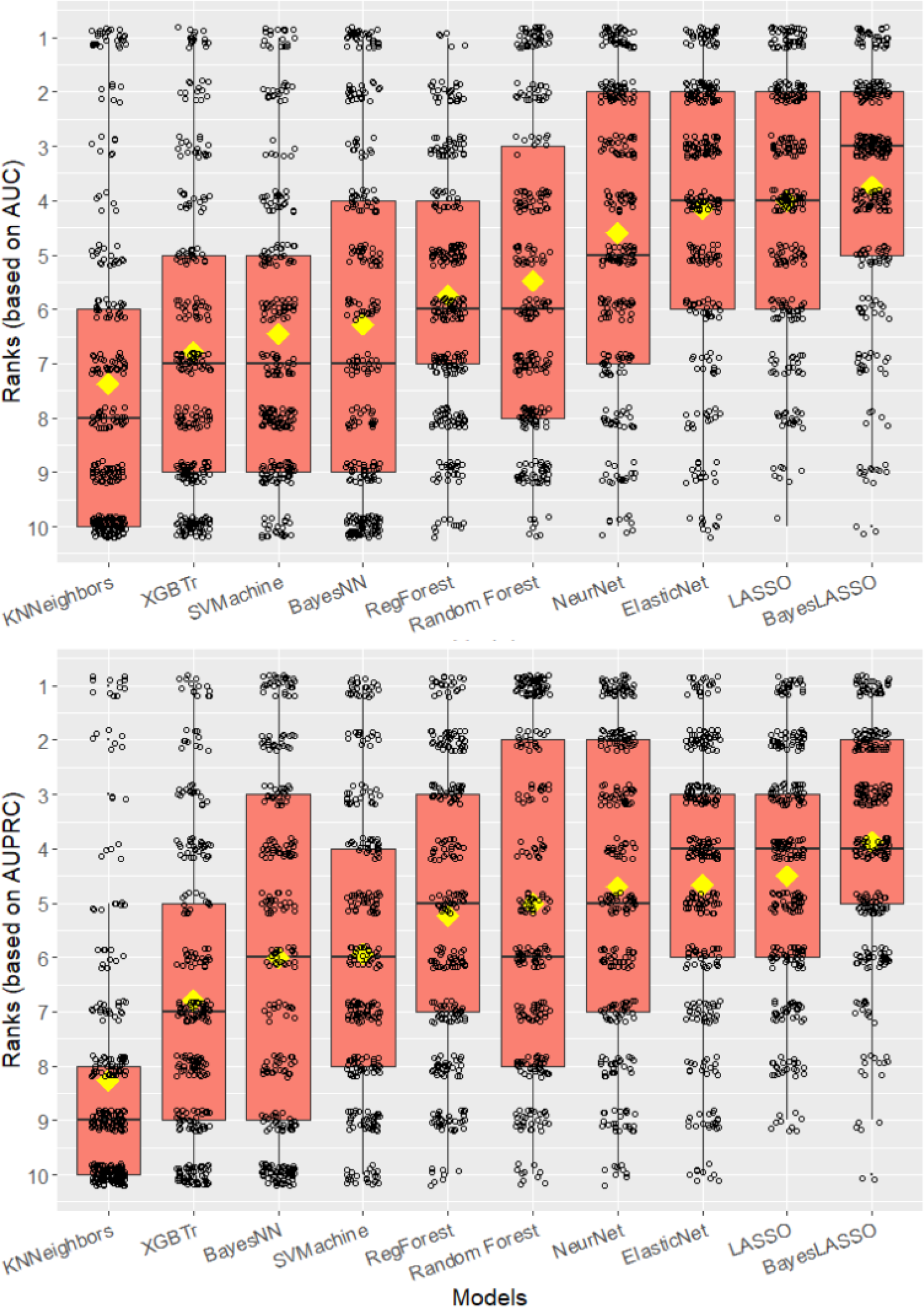
A heat map of the ranks for each algorithm across 300 separate iterations. Obtaining a rank of 1 means the algorithm had the highest AUC (top) or AUPRC (bottom) for a particular iteration. The line on each box marks the median value of the ranks. The yellow diamond locates the mean of the ranks. The models are ordered based on their mean rank.

### Interpretability

Shapley values have emerged as a powerful tool for interpreting ML models. These values offer a mathematically rigorous method to assign a proportional contribution to each feature in a predictive model. In toxicity prediction, Shapley values enable us to understand the influence of each feature on the predictions of each model [21].

Shapley values quantify the contribution of each feature by considering all their possible combinations and their corresponding impact on the prediction. This involves calculating the average marginal contribution of a feature across all possible feature combinations. This process ensures an unbiased assessment of feature importance. They are the only method that satisfies symmetry, null effect, and additivity, making them unique in the landscape of interpretability techniques [22].

In toxicity prediction, where decisions can significantly impact patient outcomes, understanding why a model makes a prediction is critical. In our application, Shapley values convert the feature values of an individual patient into values referred to as “phi”. The phi values are model-specific, and they serve as quantitative indicators of the extent to which each feature alters the toxicity risk of a patient.

In our GUI, we have incorporated the computation of Shapley values for all ten algorithms. This component enhances the transparency of the models and aids in comparative analysis, helping researchers and practitioners identify the most influential factors in toxicity prediction. By integrating Shapley values, our tool ensures that the predictions are interpretable and clinically actionable.

### Statistical Analysis

A p-value < 0.05 was considered statistically significant. The R programming language (version 4.1.1, 2021-08-10, R Core Team, Vienna, Austria) was used to conduct all the analyses in this study [15]. To construct and assess all the different ML algorithms, we utilized the open-source R package caret (version 6.0.90), a tool that has demonstrated its ability to deliver competitive results [16]. The paired Wilcoxon signed-rank test was used to compare the performance of the different model types by comparing each model in paired ranks for each iteration of model training and analysis.

## RESULTS

The model comparison process involves training toxicity models based on 10 distinct algorithms using the same training set for a single toxicity dataset. Specific metrics are then calculated for each model using the same test set for comparison. This procedure is repeated 100 times for the same dataset, randomly changing the training and test set each iteration. In our study, the methodology was applied to three separate cases of radiation-induced toxicities, resulting in 300 comparative evaluations. The total computation time was approximately 3 days on a 4-Core Intel i7-8665U Processor with 32 GB of RAM. The full analysis was performed using the GUI. All figures shown in the results are part of the output generated by this interface.

### All Datasets

Figure 2 displays the distribution of algorithm rankings based on the average AUC and AUPRC (3 toxicities × 100 repetitions = 300 data points per algorithm). Figure 3 illustrates pairwise comparisons for each pair of models (300 comparisons per pair).

**Figure 3.**
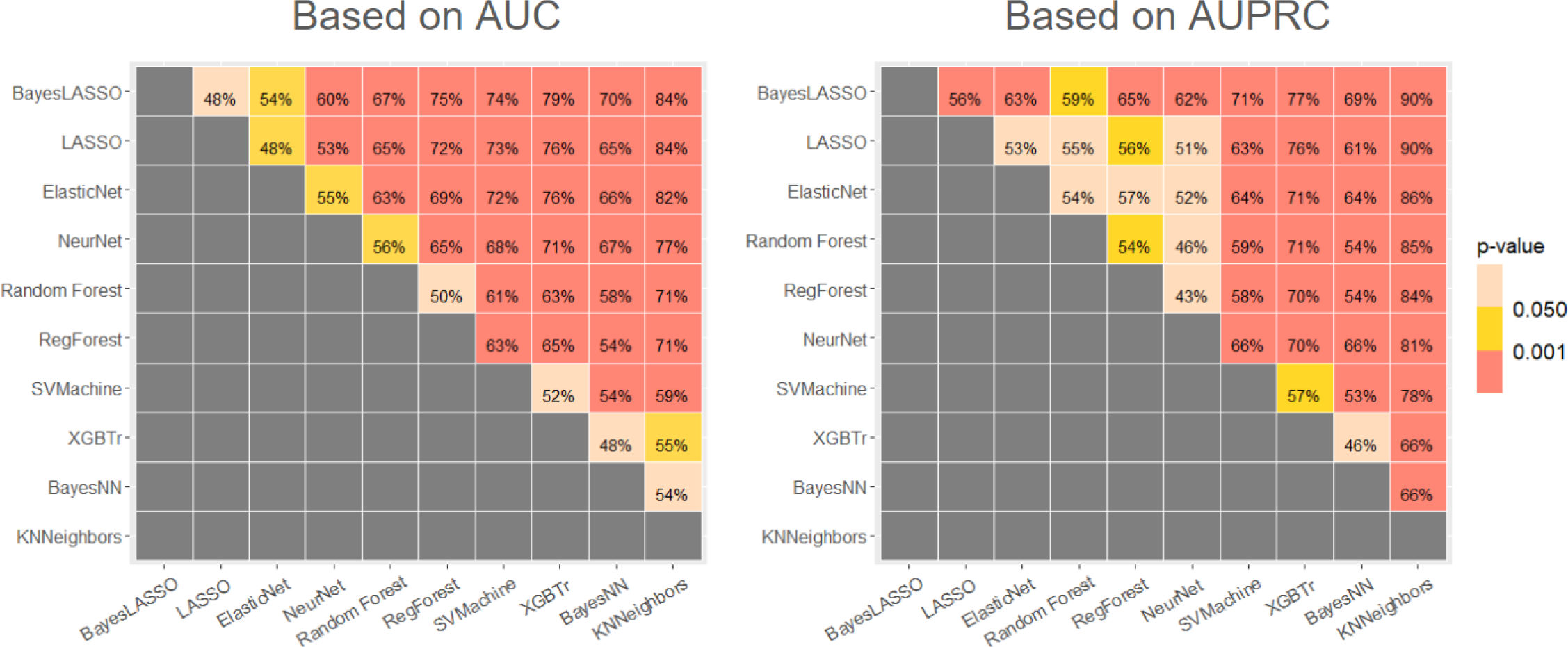
A plotted table showing pairwise comparisons among the algorithms. The numbers inside the cells represent the frequency in percentage (out of 300 comparisons) that the models in the vertical axis yielded a superior AUC (left) or AUPRC (right) than the models in the horizontal axis. The fill color corresponds to the p-value for the separation of the AUC (left) or AUPRC (right) distributions of the pairs. The gray cells represent untested pairs due to redundancy.

The Bayesian-LASSO, LASSO, and elastic net models exhibited the highest median rank (based on AUPRC) (Fig. 2). When using the Wilcoxon signed-rank test for AUPRC pairwise comparisons, Bayesian-LASSO was significantly better than all other algorithms (Fig. 3).

### Dataset Dependence

Random forest, the neural network, and LASSO achieved superior AUPRC values for the GIT, RP, and RE datasets respectively (Fig. 4). This shows that the choice of algorithm may depend on the dataset of interest.

**Figure 4.**
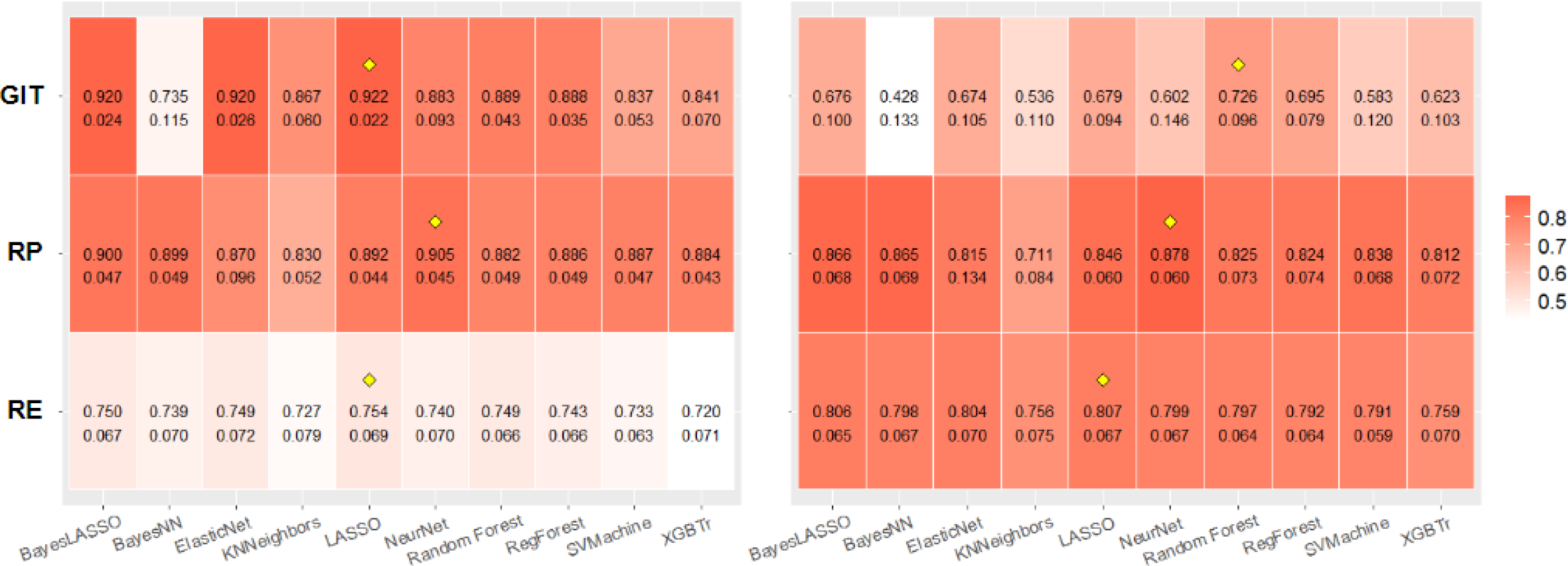
A plotted table showing the average AUC (left) and AUPRC (right) of the models for each dataset and the standard deviation of each distribution (corresponding to the 100 iterations per dataset). The yellow diamonds highlight the models which were superior for each individual cohort.

### Algorithm Selection Criteria

In Table 2, the mean AUPRCs, averaged over all 100 repetitions, are shown for random model selection and for dataset specific model selection for each dataset according to the type of toxicity. The dataset specific model selection results demonstrate an improvement of 0.066 for mean AUPRC. Comparing this method to random selection using the Wilcoxon signed-rank test shows that the improvement in AUPRC is statistically significant.

**Table 1.**
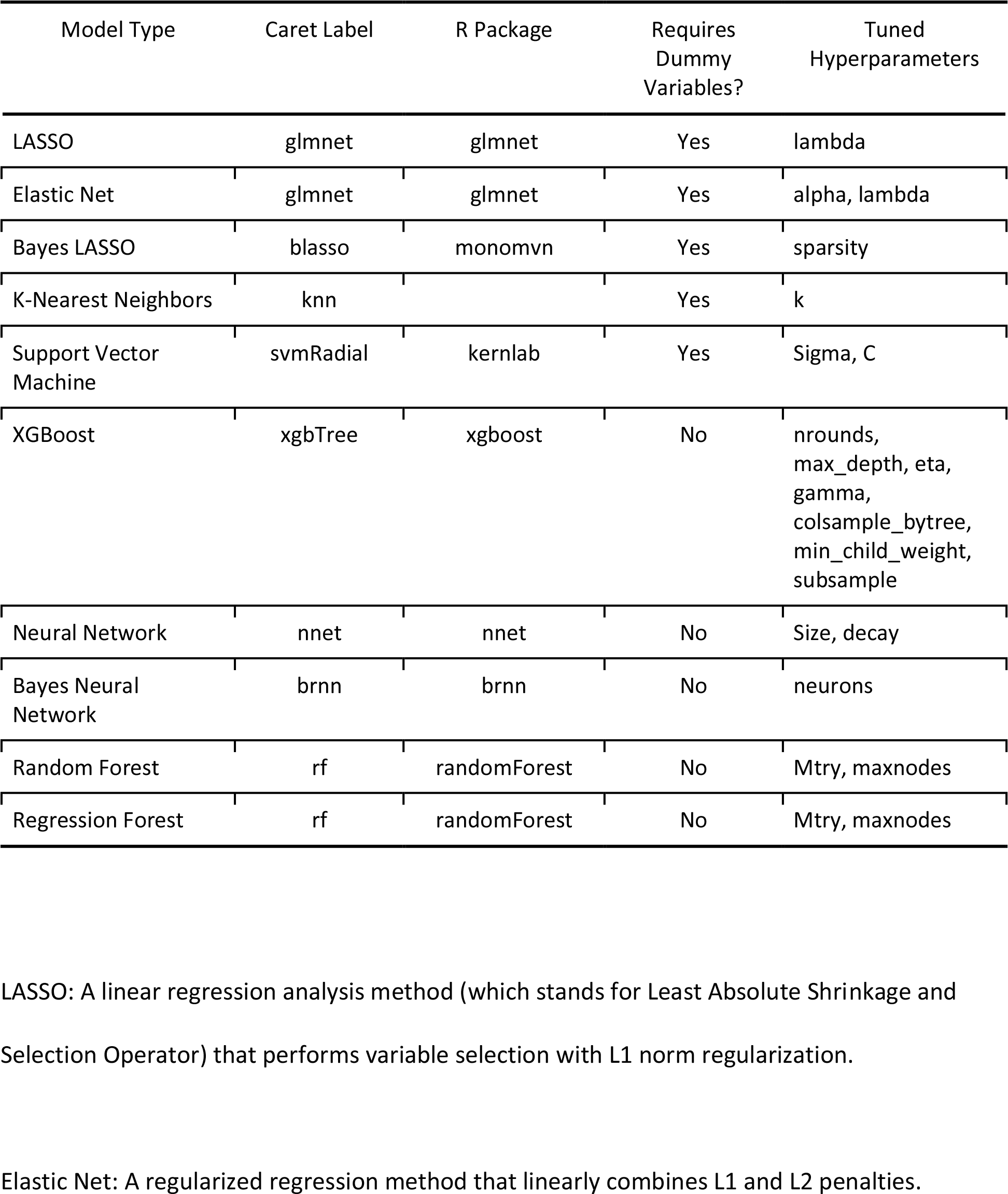
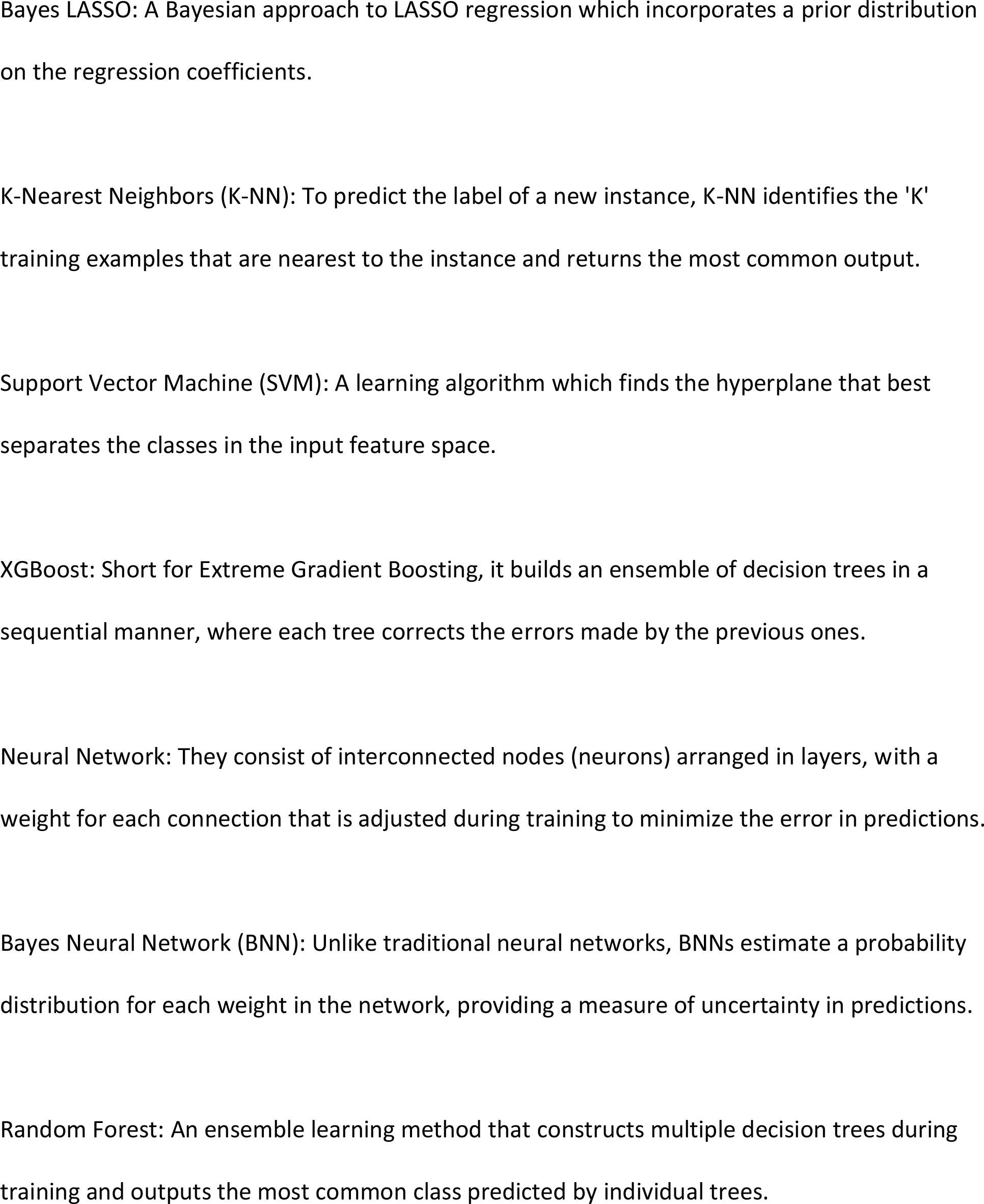
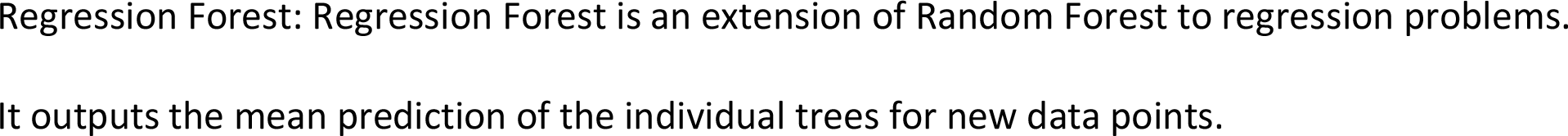
Basic characteristics of the algorithms.

| Model Type | Caret Label | R Package | Requires Dummy Variables? | Tuned Hyperparameters |
| --- | --- | --- | --- | --- |
| LASSO | glmnet | glmnet | Yes | lambda |
| Elastic Net | glmnet | glmnet | Yes | alpha, lambda |
| Bayes LASSO | blasso | monomvn | Yes | sparsity |
| K-Nearest Neighbors | knn |  | Yes | k |
| Support Vector Machine | svmRadial | kernlab | Yes | Sigma, C |
| XGBoost | xgbTree | xgboost | No | nrounds, max_depth, eta, gamma, colsample_bytree, min_child_weight, subsample |
| Neural Network | nnet | nnet | No | Size, decay |
| Bayes Neural Network | brnn | brnn | No | neurons |
| Random Forest | rf | randomForest | No | Mtry, maxnodes |
| Regression Forest | rf | randomForest | No | Mtry, maxnodes |

LASSO: A linear regression analysis method (which stands for Least Absolute Shrinkage and Selection Operator) that performs variable selection with L1 norm regularization.
Elastic Net: A regularized regression method that linearly combines L1 and L2 penalties.
Bayes LASSO: A Bayesian approach to LASSO regression which incorporates a prior distribution on the regression coefficients.
K-Nearest Neighbors (K-NN): To predict the label of a new instance, K-NN identifies the 'K' training examples that are nearest to the instance and returns the most common output.
Support Vector Machine (SVM): A learning algorithm which finds the hyperplane that best separates the classes in the input feature space.
XGBoost: Short for Extreme Gradient Boosting, it builds an ensemble of decision trees in a sequential manner, where each tree corrects the errors made by the previous ones.
Neural Network: They consist of interconnected nodes (neurons) arranged in layers, with a weight for each connection that is adjusted during training to minimize the error in predictions.
Bayes Neural Network (BNN): Unlike traditional neural networks, BNNs estimate a probability distribution for each weight in the network, providing a measure of uncertainty in predictions.
Random Forest: An ensemble learning method that constructs multiple decision trees during training and outputs the most common class predicted by individual trees.
Regression Forest: Regression Forest is an extension of Random Forest to regression problems. It outputs the mean prediction of the individual trees for new data points.

**Table 2.** In this table two different model selection criteria are compared. The random algorithm method picks an algorithm at random for each iteration and calculates its AUPRC. For the dataset-specific algorithm method, the best algorithm is selected based on the performance of the models during the training phase of all the iterations. An average AUPRC increase of 0.066 is obtained by using the latter method.

| Dataset | Random Algorithm | Dataset-specific Algorithm |  |
| --- | --- | --- | --- |
|  | AUPRC | AUPRC |  |
|  | Mean | Mean | Increase |
| SCCA | 0.602 | 0.726 | 0.124 |
| NSCLC (RE) | 0.788 | 0.807 | 0.019 |
| NSCLC (RP) | 0.822 | 0.878 | 0.056 |
| Mean |  |  | 0.066 |

### Shapley Values

The models were interpreted using Shapley values. The most important features are shown in Figure 5. Note that for GIT and RP, the best feature was not always the same one. For all 3 toxicities, the combination of best features varies by model type.

**Figure 5.**
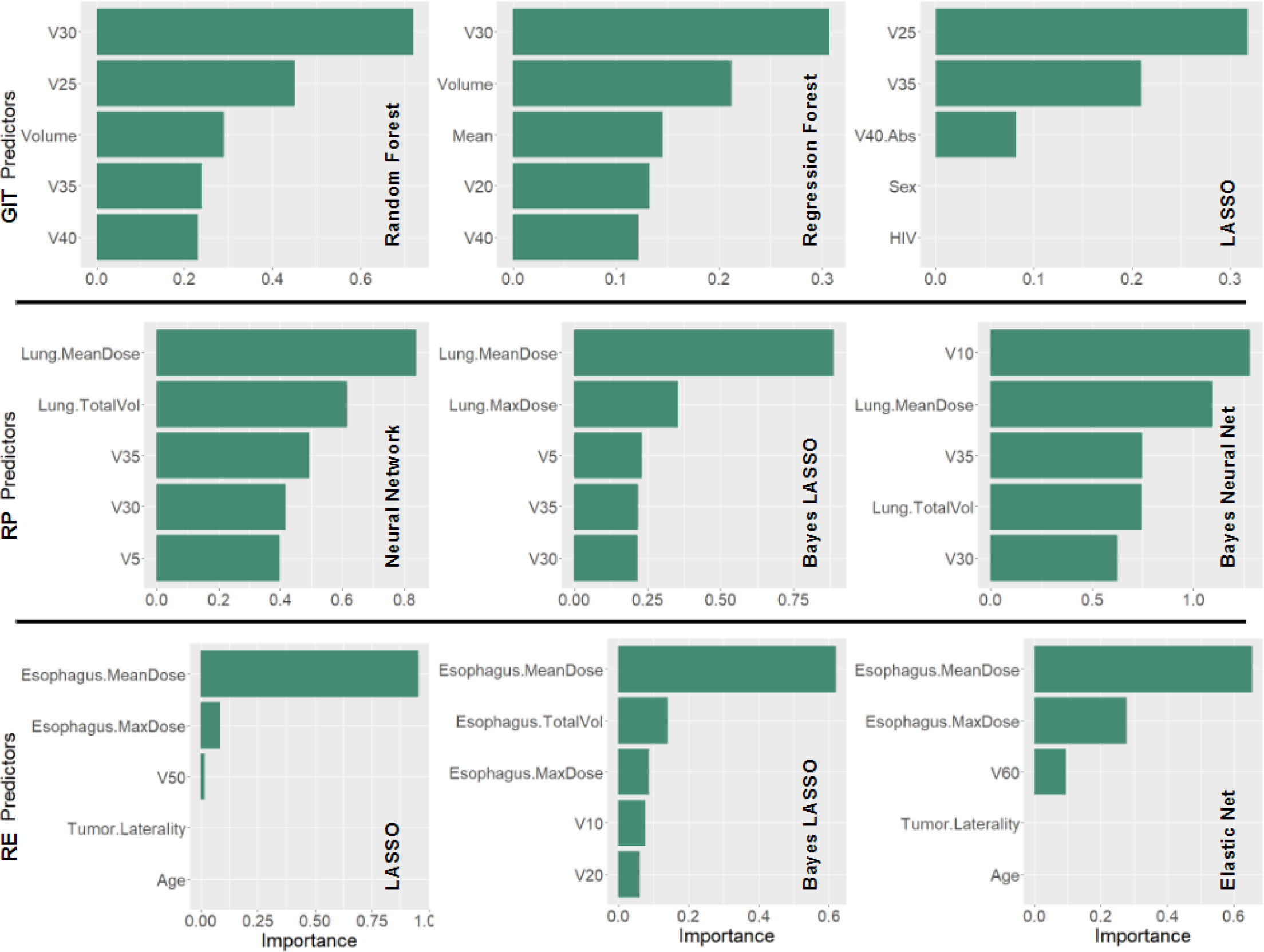
The 5 best feature for the best 3 models for each toxicity. The top, middle, and bottom rows contain information for GI, RP, and RE toxicities, respectively. The name of the model is shown on the bottom right corner of each plot. The importance was calculated based on how much each feature impacts the predicted probability of a patient developing a toxicity according to each model.

**Figure 6.**
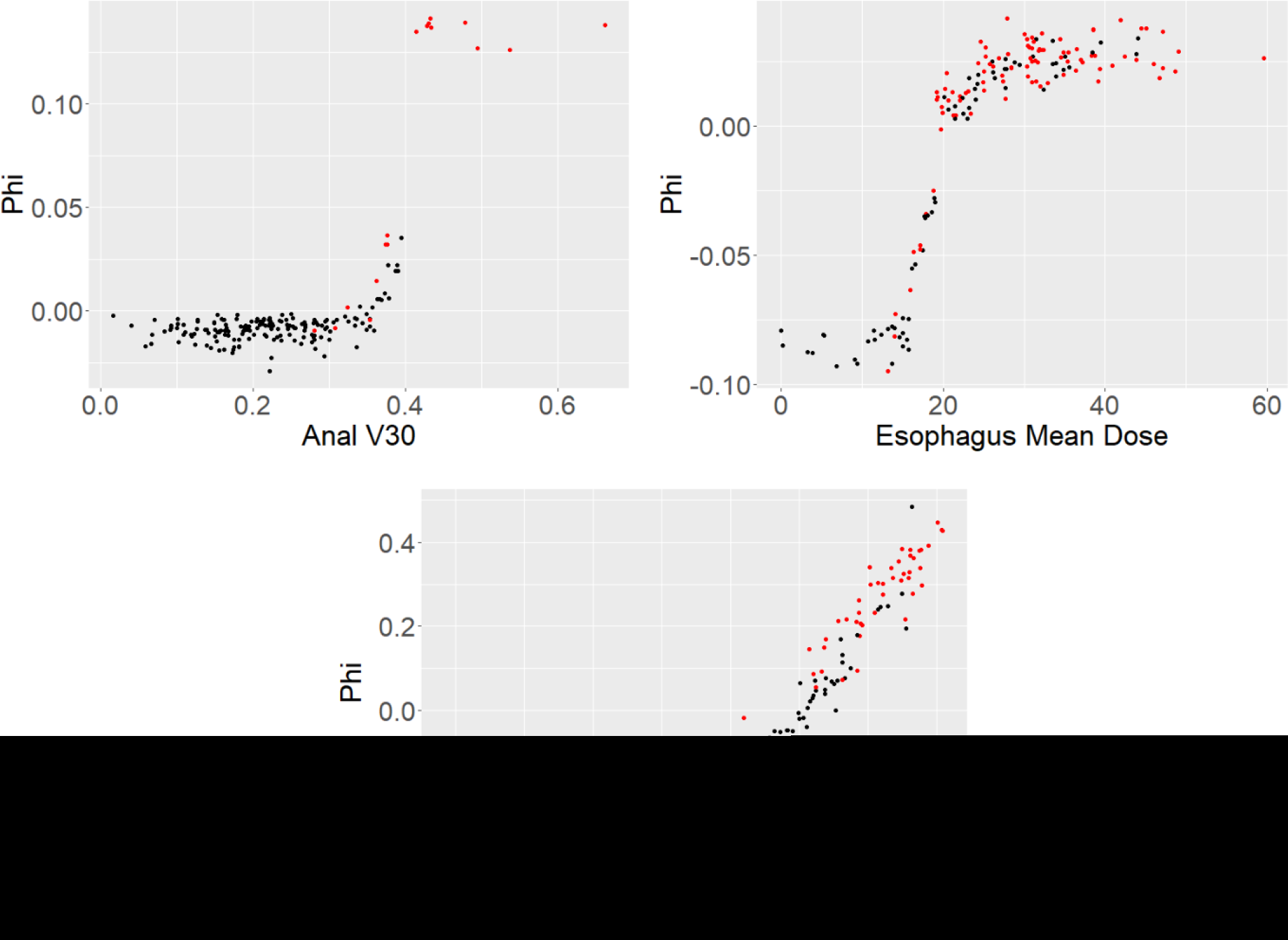
Feature value plotted against phi (the approximate change in predicted probability of toxicity due to the given feature value) for the most important features of the 3 toxicities. Black dots represent patients who did not develop the toxicity while red dots represent those who did.

By isolating the most important features, it is possible to calculate their individual importance as it varies across patients (Fig. 6). We can then identify regions for these features where small modifications may have an appreciable impact in the probability of toxicity.

## DISCUSSION

In our study, we trained and tested toxicity models using 10 distinct algorithms, conducting 100 comparisons of their performance for each of our three radiation-induced toxicities. We found that no algorithm was superior across all the different toxicities, emphasizing the importance of dataset-dependent algorithm selection. However, our findings underscore the superior predictions of the Bayesian-LASSO algorithm which outperforms other models when averaged across the analyzed toxicities. This result is noteworthy, as it highlights the robustness of the Bayesian-LASSO in diverse scenarios, suggesting that it should be a preferred choice in clinical applications for toxicity prediction. This is of particular interest as the formulation of this algorithm is relatively simple to interpret compared to other more complex, black-box type algorithms [23].

While Bayesian-LASSO generally excelled, random forest, the neural network, and LASSO showed superior performance for individual datasets (GIT, RP, and RE, respectively). This underlines the importance of comparing multiple algorithms when training models for toxicity prediction with radiation therapy datasets.

When comparing dataset specific model selection to opting for a random algorithm, the reported improvement in mean AUPRC was 0.066. If this is compared to the worst-case scenario instead (selecting the lowest performing model), the average AUPRC improvement could be as high as 0.125. Superior predictive capabilities mean clinicians can more reliably identify patients at higher risk for developing toxicities. This enables preemptive interventions, such as dose adjustments or the implementation of supportive care strategies, and personalization of treatment plans to better meet individual patient needs [24,25]. Together with the interpretability offered by Shapley values, these models can show a physician that the clinicopathological factors of a particular NSCLC patient increase their risk of toxicity, suggesting that they could benefit from a decrease in mean lung dose. Moreover, the plot relating phi to the mean lung dose illustrates the impact of this change on the toxicity risk, which helps guide clinical decision making. This aligns with the broader goals of precision medicine in oncology [26].

The results showing that data specific model selection yields an improvement over random model selection may be explained by considering the differing strengths of these algorithms. Some of these are better at modeling non-linear effects, while others are better at extrapolating and interpolating data [27]. It is then beneficial to have a systematic method for modeling any data utilizing all the algorithms mentioned in this study and comparing their predictive performance. This is the purpose of our graphical user interface.

Each separate algorithm comes with its own complexities. Comparing the performance of different algorithms on a particular dataset can become considerably time consuming for a researcher [9, 28]. The GUI developed here allows for an automatic, interactive, end-to-end multi-algorithm analysis of any radiation therapy dataset, while providing mathematically robust model interpretability through the use of Shapley values.

Our analysis and the developed GUI have significant potential for integration into heuristic clinical decision support systems [29]. By providing interpretable and accurate toxicity predictions, our models could enhance the decision-making process in radiation therapy. The use of Shapley values adds a layer of interpretability that is vital for clinical acceptance and application [30]. Clinicians can leverage these insights to understand the key factors influencing toxicity risk, allowing for more informed and personalized patient care strategies [31]. This ability to interpret the predictions given by an algorithm is critical for its credibility and clinical acceptability, and so selecting a model based on the most transparent interpretation may be desirable [32,33].

There were some limitations to our study. The analysis for all datasets was performed retrospectively, which could result in selection bias. Our work includes three toxicities, two of which are from the same treatment site (lung), and all of which are from the same institution. While this does not represent a comprehensive sample of treatment outcome datasets studied in the field of radiotherapy, the results do serve as evidence for the dataset dependence of the ML algorithms. Additionally, we did not consider the expertise of the investigator into account. Considerable expertise on a specific algorithm could justify its selection, as it has been shown that the impact of hyperparameter tuning can surpass that of algorithm selection [34].

Nonetheless, comparing those results to the performance of the top algorithms included in this study is still advisable. It should also be noted that the full comparative analysis of the different algorithms takes a considerable amount of time (approximately one day per toxicity for a dataset of about 300 patients). While this may not be an issue for a big institution, smaller clinics may not have the resources to allot this much computational time.

Given the size and variety of the data studied in this work, expanding the range of toxicities and incorporating a variety of future clinical datasets from different institutions could enhance the utility and applicability of these models. The patterns that the data on Figure 6 follow offers another approach to build upon this work. Fitting the points with carefully selected functions for all the feature plots of a dataset may allow for the creation of nomograms or other heuristic tools for different toxicities [35]. The performance of these functions can then be assessed and compared to the underlying model. The development of this GUI is meant to serve as a starting point for an evolving tool with the goal to slowly accumulate radiation therapy data to improve the overall reliability of outcome prediction models.

## CONCLUSION

We have modeled treatment outcomes across three radiation-induced toxicities utilizing ten diverse algorithms using the open-source software R, interfaced with the package caret.

Our findings provide evidence that the Bayesian-LASSO logistic regression algorithm yields superior discriminative performance on average across the datasets. However, when comparing performance within individual datasets, random forest, the neural network, and LASSO achieved the best ranks. These results demonstrate that model performance can be dataset specific, and an informed algorithm selection based on training phase performance can improve the final predictive ability. A graphical user interface provides a systematic method that automates multi-algorithm comparisons of outcome prediction models, generates visual results, and offers mathematically sound interpretations of the models.

## Data Availability

Research data are stored in an institutional repository. Anonymized data will be shared, following a 12 month embargo after the date of publication, upon request to the corresponding author and once a data transfer agreement is reached between the requestors institution and MD Anderson Cancer Center.

