## Supplemental Figures for "Performance comparison of ten state-of-the-art machine learning algorithms for outcome prediction modeling of radiation-induced toxicity"

**Supplementary Materials**

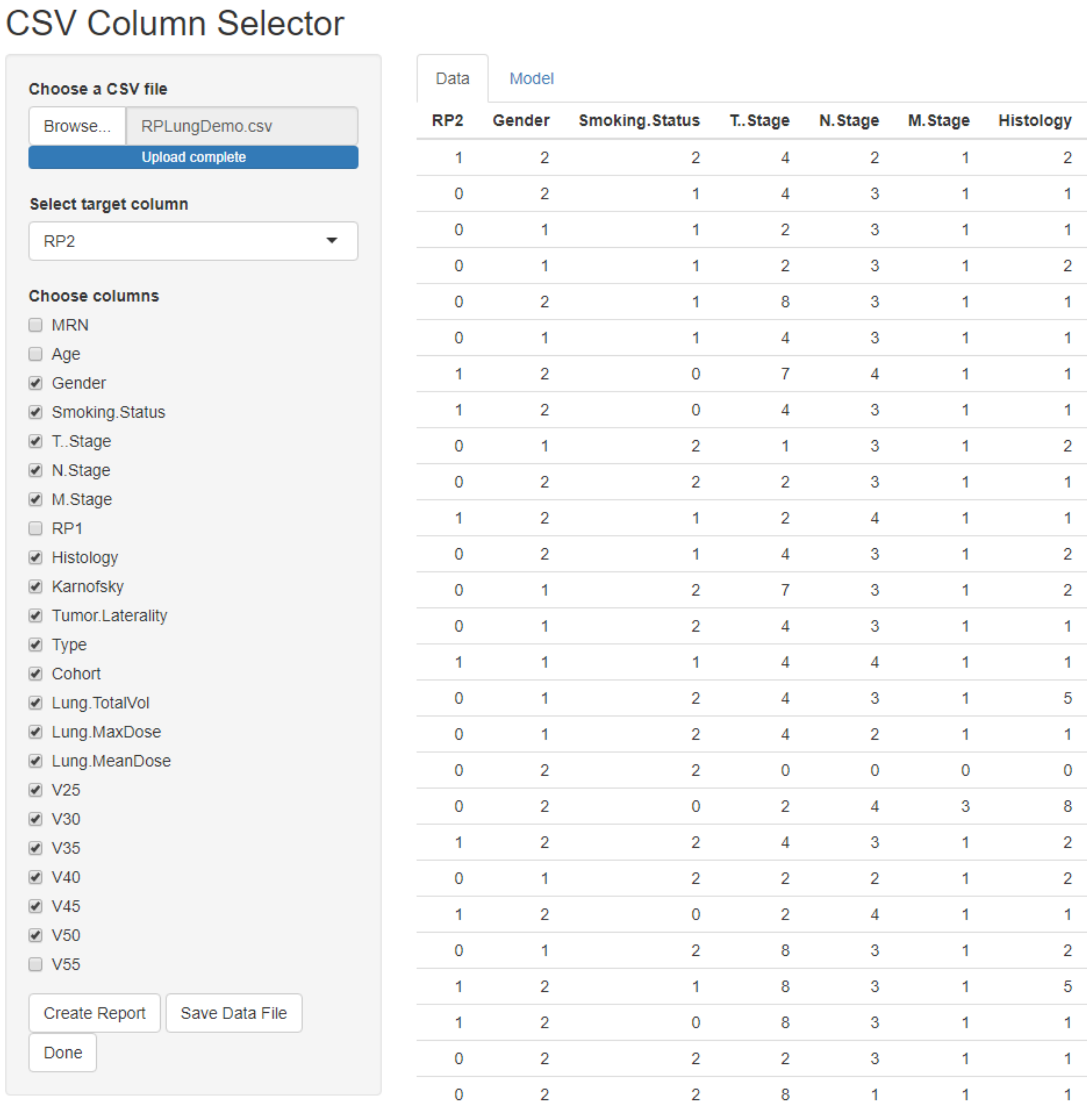


**Suppl. Figure S1.** The first tab of the graphical user interface. The “Browser” button on the top left corner of the figure was used to find the data file in its specified directory. All the features in the file are then populated inside the grey column, each with a checkbox for the user to select the ones which should be used. A drop-down menu is then provided for the user to select the column corresponding to the outcome of interest. A preview of the data is seen on the right side of the page. A button labelled “Create Report” is available to generate a document for visual exploratory data analysis (if desired). The button labelled “Save Data File” allows the user to save the data with only the selected features. The button labelled “Done” takes the user to the next tab (labelled “Model”).

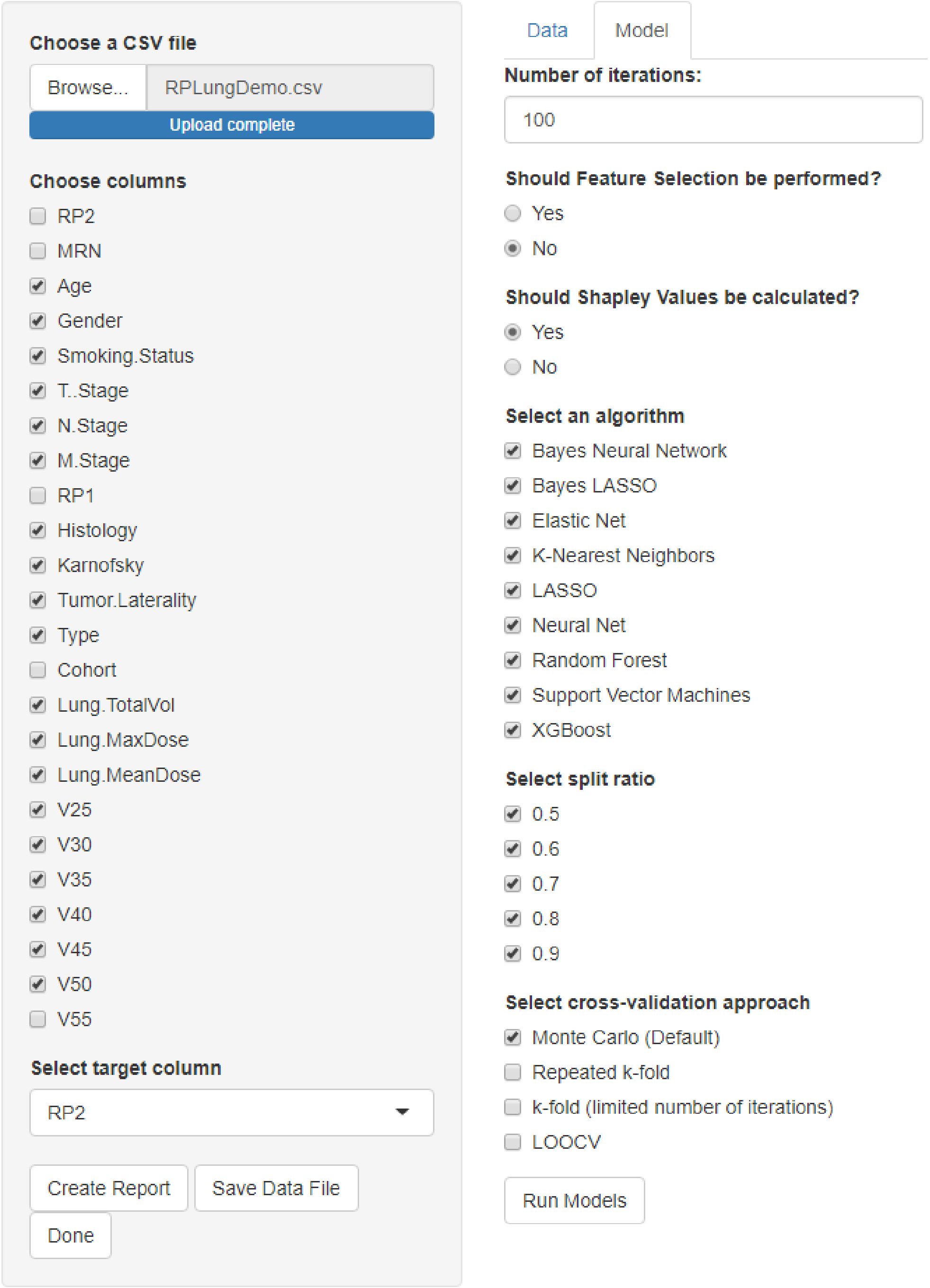


**Suppl. Figure S2**. This is the second tab of the graphical user interface. The user has the option to select the number of iterations for model comparison, whether automatic feature selection should be performed, whether the Shapley values should be calculated, the algorithms that should be trained, the split ratios to try, and the cross-validation approach that should be used. Default values for all of these are provided for convenience. Pressing the button labelled “Run Models” at the bottom right side of the figure will initiate the code.

**Suppl. Table S1.** Number of patients for different demographic, clinical, and pathological characteristics grouped into GI toxicity grades. The p-values from the univariate analysis (performed for the two different toxicity classes) for each of the shown covariates is included.

| Characteristic | | Total | Grade 0 | Grade 1 | Grade 2 | Grade 3 | Grade 4 | p-value |
|---|---|---|---|---|---|---|---|---|
| **n** | | 246 | 9 | 116 | 96 | 24 | 1 | |
| **Age (yrs)** | | | | | | | | |
| | Mean | 59.5 (34-88) | 60.2 | 59.5 | 60.0 | 62.1 | 57.9 | 0.450 |
| **Sex** | | | | | | | | |
| | Male | 57 | 2 | 35 | 19 | 1 | 0 | 0.003 |
| | Female | 189 | 7 | 81 | 77 | 23 | 1 | |
| **Smoking history** | | | | | | | | |
| | Non-smoker | 133 | 6 | 66 | 47 | 13 | 1 | 0.962 |
| | Prior smoker | 80 | 2 | 34 | 37 | 7 | 0 | |
| | Active smoker | 33 | 1 | 16 | 12 | 4 | 0 | |
| **Prior surgery** | | | | | | | | |
| | Gross total resection | 44 | 3 | 22 | 14 | 4 | 1 | 0.793 |
| | Subtotal resection | 27 | 0 | 9 | 15 | 3 | 0 | |
| | Biopsy only | 175 | 6 | 85 | 67 | 17 | 0 | |
| **HIV** | | | | | | | | |
| | Yes | 11 | 1 | 6 | 4 | 0 | 0 | 0.376 |
| | No | 235 | 8 | 110 | 92 | 24 | 1 | |
| **T Stage** | | | | | | | | |
| | 1 | 43 | 2 | 26 | 12 | 3 | 0 | 0.165 |
| | 2 | 117 | 4 | 51 | 49 | 12 | 1 | |
| | 3 | 54 | 3 | 25 | 23 | 3 | 0 | |
| | 4 | 32 | 0 | 14 | 12 | 6 | 0 | |
| **N Stage** | | | | | | | | |
| | 0 | 114 | 4 | 59 | 42 | 8 | 1 | 0.428 |
| | 1 | 132 | 5 | 57 | 54 | 16 | 0 | |
| **M Stage** | | | | | | | | |
| | No Metastasis | 238 | 9 | 114 | 92 | 22 | 1 | 0.169 |
| | Metastasis | 8 | 0 | 2 | 4 | 2 | 0 | |
| **Histology** | | | | | | | | |
| | Well differentiated | 15 | 2 | 7 | 4 | 2 | 0 | 0.754 |
| | Moderately differentiated | 109 | 2 | 60 | 38 | 9 | 0 | |
| | Poorly differentiated | 112 | 5 | 44 | 49 | 13 | 1 | |
| | Unknown | 10 | 0 | 5 | 5 | 0 | 0 | |
| **Pathology** | | | | | | | | |
| | SCCA | 205 | 6 | 100 | 80 | 18 | 1 | 0.233 |
| | SCCA/basaloid features | 34 | 3 | 14 | 11 | 6 | 0 | |
| | SCCA/mixed histology | 7 | 0 | 2 | 5 | 0 | 0 | |
| **Inguinal Nodes** | | | | | | | | |
| | Yes | 62 | 2 | 24 | 25 | 11 | 0 | 0.057 |
| | No | 184 | 7 | 92 | 71 | 13 | 1 | |
| **Tumor size (cm)** | | | | | | | | |
| | Mean | 3.7 (0.5-10) | 3.6 | 3.7 | 3.8 | 3.6 | 2.1 | 0.911 |
| **Tumor location** | | | | | | | | |
| | Anal canal | 276 | 8 | 104 | 93 | 23 | 1 | 0.705 |
| | Perianal region | 20 | 1 | 12 | 3 | 1 | 0 | |
| **Chemotherapy Sequence** | | | | | | | | |
| | Concurrent | 235 | 9 | 111 | 95 | 19 | 1 | <0.001 |
| | Sequential | 6 | 0 | 2 | 0 | 4 | 0 | |
| | Other | 5 | 0 | 3 | 1 | 1 | 0 | |
| **Agents** | | | | | | | | |
| | Cisplatin/5-FU | 185 | 3 | 89 | 77 | 15 | 1 | 0.142 |
| | MMC/5-FU | 33 | 3 | 12 | 11 | 7 | 0 | |
| | Oxali/cape | 8 | 0 | 5 | 2 | 1 | 0 | |
| | 5-FU only | 9 | 1 | 5 | 3 | 0 | 0 | |
| | Cape/cisplatin | 5 | 2 | 0 | 2 | 1 | 0 | |
| | Cape/MMC | 2 | 0 | 2 | 0 | 0 | 0 | |
| **RT Technique** | | | | | | | | |
| | IMRT | 143 | 7 | 71 | 48 | 16 | 1 | 0.248 |
| | VMAT | 103 | 2 | 45 | 48 | 8 | 0 | |
| **Fractions** | | | | | | | | |
| | Median | 27 (18-30) | 27 | 27 | 27 | 27 | 27 | 0.965 |
| **Rx Dose** | | | | | | | | |
| | Median | 5500 (4500-6000) | 5400 | 5400 | 5400 | 5400 | 5400 | 0.909 |

Abbreviations: n, number of patients; yrs, years; HIV, human immunodeficiency virus; T Stage, tumor size; N Stage, number of nearby lymph nodes that have cancer; M Stage, metastasis status; SCCA, squamous cell carcinoma of the anus; 5-FU, 5-fluorouracil; MMC, mitomycin-C; Oxali, oxaliplatin; Cape, capecitabine; RT, radiotherapy; IMRT, intensity-modulated radiation therapy; VMAT, volumetric modulated arc therapy; Rx, prescribed dose

**Supplementary Table S2**. Patient clinical and pathological characteristics grouped according to radiation pneumonitis severity (symptomatic RP, Grade ≥2; asymptomatic RP, Grade <2).

| Characteristic | | Total | RP < 2 | RP > 2 | p-value |
|---|---|---|---|---|---|
| Patients (n) | | 232 | 164 | 68 | |
| Age (yrs) | | | | | |
| | Mean | 65.3 (33-88) | 65.4 | 65.0 | 0.77 |
| Gender | | | | | |
| | Male | 126 | 91 | 35 | 0.68 |
| | Female | 106 | 73 | 33 | |
| Smoking History | | | | | |
| | Non-smoker | 19 | 9 | 10 | 0.03 |
| | Prior smoker | 165 | 123 | 42 | |
| | Active smoker | 48 | 32 | 16 | |
| Karnofsky Score | | | | | |
| | <=60 | 9 | 8 | 1 | 0.23 |
| | 70-80 | 155 | 107 | 48 | |
| | 90-100 | 68 | 49 | 19 | |
| T Stage | | | | | |
| | T0/T1 | 41 | 31 | 10 | 0.21 |
| | T2 | 85 | 54 | 31 | |
| | T3 | 43 | 28 | 15 | |
| | T4 | 63 | 51 | 12 | |
| N Stage | | | | | |
| | N0 | 22 | 20 | 2 | 0.05 |
| | N1 | 24 | 18 | 6 | |
| | N2 | 122 | 88 | 34 | |
| | N3 | 64 | 38 | 26 | |
| M Stage | | | | | |
| | M0 | 223 | 157 | 66 | 0.45 |
| | M1 | 9 | 7 | 2 | |
| Histology | | | | | |
| | Adeno | 124 | 84 | 40 | 0.47 |
| | Squamous | 76 | 57 | 19 | |
| | Poorly differentiated NSCLC | 1 | 1 | 0 | |
| | NSC NOS | 5 | 5 | 0 | |
| | Mixed NSC | 1 | 0 | 1 | |
| | Large Cell | 2 | 2 | 0 | |
| | Carcinoid | 2 | 1 | 1 | |
| | Other | 17 | 11 | 6 | |
| | Unknown | 4 | 3 | 1 | |
| Induction Chemo | | | | | |
| | Yes | 29 | 21 | 8 | 0.83 |
| | No | 203 | 143 | 60 | |
| Concurrent Chemo | | | | | |
| | Yes | 226 | 158 | 68 | 0.25 |
| | No | 6 | 6 | 0 | |
| Tumor Location | | | | | |
| | RUL | 97 | 64 | 33 | 0.08 |
| | RML | 37 | 28 | 9 | |
| | RLL | 24 | 13 | 11 | |
| | LUL | 43 | 34 | 9 | |
| | LLL | 22 | 16 | 6 | |
| | Mediastinum | 9 | 9 | 0 | |
| GTV Volume | | | | | |
| | Mean | 129.57 (0 - 2017.15) | 123.55 | 146.67 | <0.001 |

*Abbreviations: n, number of patients; age, years; T Stage, tumor size; N Stage, number of nearby lymph nodes that have cancer; M Stage, metastasis status; Chemo, chemotherapy; GTV, gross tumor volume; NSCLC, non-small cell lung cancer; NOS, not otherwise specified; RUL, right upper lung; RML, right middle lung; RLL, right lower lung; LUL, left upper lung; LLL, left lower lung.*

**Supplementary Table S3**. Patient clinical and pathological characteristics grouped according to radiation esophagitis severity (symptomatic RE, Grade ≥2; asymptomatic RE, Grade <2).

| Characteristic | | Total | RE < 2 | RE > 2 | p-value |
|---|---|---|---|---|---|
| Patients (n) | | 232 | 98 | 134 | |
| Age (yrs) | | | | | |
| | Mean | 65.3 (33-88) | 66.4 | 64.5 | 0.11 |
| Gender | | | | | |
| | Male | 126 | 58 | 68 | 0.25 |
| | Female | 106 | 40 | 66 | |
| Smoking History | | | | | |
| | Non-smoker | 19 | 7 | 12 | 0.34 |
| | Prior smoker | 165 | 75 | 90 | |
| | Active smoker | 48 | 16 | 32 | |
| Karnofsky Score | | | | | |
| | <=60 | 9 | 8 | 1 | 0.45 |
| | 70-80 | 155 | 61 | 94 | |
| | 90-100 | 68 | 29 | 39 | |
| T Stage | | | | | |
| | T0/T1 | 41 | 14 | 27 | 0.15 |
| | T2 | 85 | 41 | 44 | |
| | T3 | 43 | 20 | 23 | |
| | T4 | 63 | 23 | 40 | |
| N Stage | | | | | |
| | N0 | 22 | 15 | 7 | 0.04 |
| | N1 | 24 | 14 | 10 | |
| | N2 | 122 | 50 | 72 | |
| | N3 | 64 | 19 | 45 | |
| M Stage | | | | | |
| | M0 | 223 | 96 | 127 | 0.27 |
| | M1 | 9 | 2 | 7 | |
| Histology | | | | | |
| | Adeno | 124 | 52 | 72 | 0.16 |
| | Squamous | 76 | 32 | 44 | |
| | Poorly differentiated NSCLC | 1 | 1 | 0 | |
| | NSC NOS | 5 | 4 | 1 | |
| | Mixed NSC | 1 | 1 | 0 | |
| | Large Cell | 2 | 1 | 1 | |
| | Carcinoid | 2 | 1 | 1 | |
| | Other | 17 | 3 | 14 | |
| | Unknown | 4 | 3 | 1 | |
| Induction Chemo | | | | | |
| | Yes | 29 | 14 | 15 | 0.62 |
| | No | 203 | 84 | 119 | |
| Concurrent Chemo | | | | | |
| | Yes | 226 | 93 | 133 | 0.10 |
| | No | 6 | 5 | 1 | |
| Tumor Location | | | | | |
| | RUL | 97 | 36 | 61 | 0.23 |
| | RML | 37 | 18 | 19 | |
| | RLL | 24 | 11 | 13 | |
| | LUL | 43 | 22 | 21 | |
| | LLL | 22 | 10 | 12 | |
| | Mediastinum | 9 | 1 | 8 | |
| GTV Volume | | | | | |
| | Mean | 129.57 (0 - 2017.15) | 120.98 | 135.85 | <0.001 |

*Abbreviations: n, number of patients; age, years; T Stage, tumor size; N Stage, number of nearby lymph nodes that have cancer; M Stage, metastasis status; Chemo, chemotherapy; GTV, gross tumor volume; NSCLC, non-small cell lung cancer; NOS, not otherwise specified; RUL, right upper lung; RML, right middle lung; RLL, right lower lung; LUL, left upper lung; LLL, left lower lung.*
